# The cost of sleeping sickness vector control in the Democratic Republic of the Congo

**DOI:** 10.1101/2024.02.02.24302172

**Authors:** Rian Snijders, Alexandra P.M. Shaw, Richard Selby, Inaki Tirados, Paul R Bessell, Alain Fukinsia, Erick Miaka, Fabrizio Tediosi, Epco Hasker, Marina Antillon

## Abstract

Gambian human African trypanosomiasis (gHAT), a neglected tropical disease caused by a parasite transmitted by tsetse flies, once inflicted over 30,000 annual cases and resulted in half a million deaths in the late twentieth century. An international gHAT control program has reduced cases to under 1,000 annually, encouraging the World Health Organization to target the elimination of gHAT transmission by 2030. This requires adopting innovative disease control approaches in foci where transmission persists. Since the last decade, case detection and treatment, the mainstay of controlling the disease, is supplemented by vector control using Tiny Targets, small insecticide-treated screens, which attract and kill tsetse. The advantages of Tiny Targets lie in their relatively low cost, easy deployment, and effectiveness.

The Democratic Republic of Congo (DRC), bearing 65% of the 799 gHAT cases reported globally in 2022, introduced Tiny Targets in 2015. This study estimates the annual cost of vector control using Tiny Targets in a health district in the DRC and identifies the main cost drivers. Economic and financial costs, collected from the provider’s perspective, were used to estimate the average cost of tsetse control expressed as cost (i) per target used, (ii) per target deployed, (iii) linear kilometre of river controlled, and (iv) square kilometres protected by vector control. Sensitivity analyses were conducted on key parameters for results robustness.

The estimated annual economic cost for protecting an area of 1,925 km² was 120,000 USD. This translates to 5.3 USD per target used each year, 11 USD per target deployed in the field, 573 USD per linear km treated, and 62 USD per km² protected. These costs in the DRC are comparable to those in other countries. The study provides valuable information for practitioners and policymakers aiding them in making rational, evidence-based decisions regarding cost-effective strategies to control gHAT.

**Author Summary:** In the fight against Gambian human African trypanosomiasis (gHAT), a devastating disease transmitted by tsetse flies, significant progress has been made through international efforts. Despite the annual cases being reduced to under 1,000, the World Health Organization aims to eliminate gHAT transmission by 2030. A key component of this strategy involves innovative approaches, such as the use of Tiny Targets – small, cost-effective, insecticide-treated screens that attract and kill tsetse flies. This study focuses on the Democratic Republic of Congo (DRC), which bears a substantial burden of gHAT cases, estimating the annual cost of vector control using Tiny Targets in a specific health district. The analysis, conducted from the provider’s perspective, reveals an annual economic cost of 120,000 USD for protecting a 1,925 km² area. This translates to 5.3 USD per target used, 11 USD per target deployed, 573 USD per linear km treated, and 62 USD per km² protected. These findings, comparable to costs in other countries, offer valuable insights for practitioners and policymakers, guiding evidence-based decisions on cost-effective strategies for gHAT control.

## Introduction

Sleeping sickness or Human African Trypanosomiasis (HAT) is a vector-borne parasitic disease that caused several major outbreaks in sub-Saharan Africa, killing thousands of people during the last epidemic of the 1990s. The disease is transmitted through the bite of an infected tsetse fly (genus *Glossina*) and is almost always lethal if left untreated [1]. This article focuses on the cost of vector control, one of the approaches to control the disease, by reducing the population of tsetse flies responsible for transmitting the parasite [2].

By 1960, colonial authorities almost eliminated HAT, previously a major public health problem affecting millions of people in Sub-Saharan Africa. This was achieved by implementing –occasionally oppressive–case-finding measures and providing effective but highly toxic treatments. Unfortunately, the disease re-emerged and peaked at the end of the 1990s, with over 30,000 new cases reported annually, causing a significant social and economic impact on the affected regions. Stepping up HAT control and surveillance efforts from the late 1990s onwards reversed this epidemiological trend, with around 6,200 cases reported in 2013 [3].

Therefore, that year, the international community declared that HAT elimination was feasible because of the sustained decrease in the disease burden, a better understanding of the disease’s epidemiology, and the prospect of improved diagnostics and treatment regimens that were less toxic than previously. The World Health Organization (WHO) set 2020 as the target date for HAT elimination as a public health problem, defined as reducing HAT incidence to fewer than 1 new case per 10,000 population in at least 90% of foci and to fewer than 2,000 cases reported globally. They also targeted 2030 as the year for disease elimination, defined as zero disease incidence [2, 4–6]. In 2020, the elimination target was largely achieved, with only 663 new HAT cases reported globally, 60% of which were identified in the Democratic Republic of Congo (DRC) [7]. In 2022, 516 or 65% of the gHAT cases reported globally were detected in the DRC [3].

Two forms of HAT exist in humans, caused by two subspecies of the parasite *Trypanosoma brucei,* namely *T. b. gambiense* infections currently responsible for over 85% of all HAT cases reported worldwide and *T. b. rhodesiense.* Both forms of HAT are targeted for elimination as a public health problem, but only gambiense HAT (gHAT) is targeted for elimination of transmission to humans as it is presumed to be an anthroponotic infection, unlike *T. b. rhodesiense* HAT, which can infect animals [7].

gHAT is the only form of HAT present in the DRC, and HAT control in this context focuses on clearing the parasite from humans [8]. This strategy is based on a multi-faceted approach, focusing mainly on early case detection and treatment [2]. HAT diagnosis is difficult because of its non-specific symptoms, the diagnostic algorithm’s complexity, and the disease’s focal distribution [1]. Therefore, an exhaustive screening strategy, even with innovative diagnostics and treatment, requires major investments in equipment, diagnostics, and human resources [2, 9]. Even though case-finding strategies have proven to be effective, there are still several foci where transmission persists, sometimes even after HAT case detection and management has been maintained for many years, most likely due to insufficient coverage of the population at risk, limited sensitivity of diagnostic methods and the disappearing awareness and expertise of medical staff [10, 11].

In the past, vector control methods, such as vegetation clearing, or insecticide spraying, and in particular, trapping, were occasionally used to control gHAT but were often considered ineffective, too expensive, or too complicated to implement in remote, resource-constrained settings [12–15]. Today, a new, more straightforward and cost-effective method has been developed to control populations of riverine tsetse that transmit *T. b. gambiense*, namely ‘Tiny Targets’. These small, impregnated screens consist of one 25cm by 25cm square blue cloth, flanked by an insecticide-impregnated mesh of the same size, deployed along the banks of rivers and water bodies where tsetse concentrate. Tsetse are attracted by their blue colour and contact the targets, picking up a lethal dose of insecticide. In 2011, Tiny Targets were introduced in Guinea and Uganda where their relative entomological and economic cost-effectiveness as compared to previous methods is discussed [16–18]. Afterwards, this relatively cheap tool was effectively deployed in several other countries (e.g., Chad, Côte d’Ivoire), achieving in all locations a decline in the tsetse population of 60-95% [19–21]. HAT transmission models estimated that at least a 72% reduction in the tsetse population is required to stop transmission and that the 2030 gHAT elimination goal would be achieved by including a moderately effective tsetse control (60% tsetse population reduction) in the overall gHAT control strategy. Therefore, vector control could be crucial in eliminating gHAT cost-effectively [18, 22].

In 2015, gHAT vector control using Tiny Targets was implemented at the health district level in the DRC for the first time. An evaluation of the impact of Tiny Targets on the *Glossina fuscipes quanzensis*, the primary tsetse vector of gHAT in the DRC observed a reduction in fly catches of more than 85% [13]. This study estimates the annual financial and economic cost of Tiny Target deployment and identifies its main cost drivers.

## Materials and Methods

### Research setting

The health system of the DRC is organized at different levels, where every province is subdivided into several health districts where a district team manages a network of health centres and a district hospital. Each health district generally covers a human population between 100,000 and 200,000 which according to national standards is subdivided into health areas of around 10,000 inhabitants each, covered by at least one integrated health centre [23].

As of 2014, a first project started in the health districts of Yasa Bonga and Mosango focussing on improving HAT control named “Integrated HAT control, a model district in DR Congo” and by the end of 2015 a second project was introduced in the same districts named “TRYP-ELIM. A demonstration project combining innovative case detection, tsetse control and IT to eliminate sleeping sickness at district level in the Democratic Republic of Congo”. These projects aimed to effectively eliminate HAT transmission within three years from a health district in the DRC through a combination of intensified systematic screening and case management of at-risk populations with vector control. In the context of this project, tsetse vector control with Tiny Targets (manufactured by Vestergaard, Lausanne, Switzerland) was implemented throughout the HAT-endemic health district of Yasa Bonga in Kwilu province, formerly Bandundu Province. Yasa Bonga is a rural health district with, in 2018, a total population of 235,696 people scattered over 305 villages in an area of 2,810 km^2^ [13, 24]. Over 45% of all HAT cases detected in the DRC were reported in the former Bandundu between 2000 and 2012, with the highest annual incidence, of 40 cases/10,000 population (208 new cases), being reported in Yasa Bonga. [25].

### Vector control with Tiny Targets in the study area

Figure 1 shows the gradual scale up of vector control with Tiny Targets in the Yasa Bonga health district along the riverbanks of some of the three main rivers (Lukula, Kafi, Inzia). In 2015, the rivers highlighted in red were treated, the following year the river to the north was included as well, and in 2017 treatment was extended to include part of the river forming the western border of the health district. Thus, by 2017, the health areas highlighted in green were covered so that almost the whole health district was protected.

**Figure 1.**
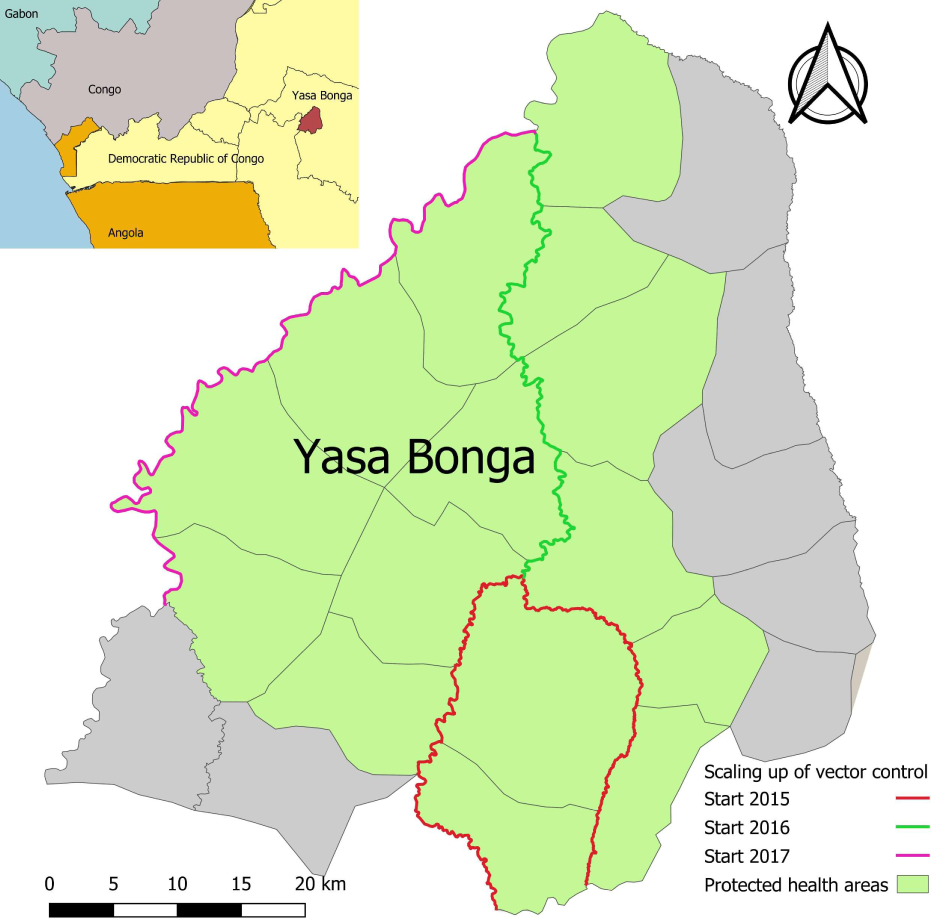
Map showing the border of Yasa Bonga and the individual health areas. Overlain is the scaling up of vector control in the health district of Yasa Bonga between 2015 and 2017 Map generated using QGIS 3.28.3 [26, 27]

The deployments, as outlined in Table 1, occurred biannually during the dry seasons (July and August and January/early February). To evaluate the impact of the Tiny Targets, the abundance of tsetse flies was assessed before and after deployments. Pyramidal traps were used to compare the number of flies captured during these periods [28]. Teams of day workers without previous experience deployed the Tiny Targets. Local recruitment and training were managed by the Programme National de Lutte contre la Trypanosomiase Humaine Africaine (PNLTHA) and with support from entomologists from the PNLTHA and the Liverpool School of Tropical Medicine (LSTM).

**Table 1.** Estimated financial cost of the full scale intervention for a 5-year period based on the costs reported between 2015 and 2017 (USD).

| Description/cost category | Total<br>Y1 | Total<br>Y2 | Total<br>Y3 | Total<br>Y4 | Total<br>Y5 | Total<br>Y1-Y5 |
| --- | --- | --- | --- | --- | --- | --- |
| Monitoring/Surveillance | 14,214 | 14,214 | 14,214 | 14,214 | 14,214 | 71,071 |
| Sensitization | 44,217 | 19,541 | 27,717 | 19,541 | 27,717 | 138,731 |
| Biannual target deployment | 57,571 | 57,571 | 57,571 | 57,571 | 57,571 | 287,855 |
| Management support costs | 52,476 | 13,305 | 13,780 | 13,755 | 19,286 | 112,601 |
| **Total cost** | **168,478** | **104,630** | **113,282** | **105,080** | **118,788** | **610,257** |
| Cost Tiny targets (1\$) | 22,622 | 22,622 | 22,622 | 22,622 | 22,622 | 113,110 |
| **Total cost with Tiny target donation** | **145,856** | **82,008** | **82,008** | **82,458** | **96,166** | **497,147** |

Teams assembled targets on-site by gluing the fabric on locally sourced wooden supports. Using traditional canoes or “pirogues”, the teams travelled down the river deploying the targets aiming for 50-meter intervals on both riverbanks preferring shorter intervals when exact 50m intervals were impossible due to unsuitable deployment or canoe docking sites, explaining a higher actual number of targets deployed than the targeted 40targets/km. The last deployment, covering the whole protected area, involved around 54 targets / linear km, resulting in 11,311 targets over 210 linear km, as illustrated in S1.Annexe I. GPS points were recorded for every target placed. Once a routine was established, the teams typically covered around 40 linear km of river weekly (i.e., 40km x 40 targets/km = 1600 targets/week) though this number increased over the years. A detailed description of the vector control intervention and the impact on the vector, can be found in Tirados *et al*. [13]. S1.Annex II shows the calendar with the different vector control activities during the study period including the number of targets deployed and linear km treated per deployment during the scale up. Awareness-building activities accompanied the vector control intervention to inform villages about sleeping sickness and tsetse control activities. A vector control management unit was set up at the provincial level after the study period in 2019, and this team manages a standardized vector control sensitization strategy in all villages where vector control measures are implemented. This strategy was used as a reference for the cost of awareness-building activities in Yasa Bonga. In the health district, 165 villages that would be in contact with the gHAT vector control were identified for sensitization. On average, the provincial vector control team biennially trained three community health care workers per village. Furthermore, a vector management control unit was set up at national level covering 11 health districts in 2 provinces. These costs were not taken into account in the main analysis but their cost impact was assessed in the sensitivity analysis. Those community healthcare workers were responsible for sensitizing other community healthcare workers and the local population. They would receive T-shirts, a picture box, a banner, and megaphones, and their work was supported with short broadcasts on local radio stations.

### Cost methodology

The cost analysis adopted a provider’s perspective focusing on costs incurred to implement HAT vector control using Tiny Targets by the public health care system, namely the PNLTHA. The study only considered costs related directly to the implementation of vector control and omitted research costs as well as costs of geostatistical modeling before the intervention to predict tsetse habitat distribution [29, 30]. A full costing approach was adopted, which was the case for other similar analyses, as similar programs are assumed to be implemented anew in different locations [17, 31, 32].

The study collected financial and economic costs at local prices between January 2015 and September 2017 from routine activities, expense reports, budgets, and discussions with experts. During this period, five target deployments took place, covering a gradually expanding area, as shown in Figure 1 with the linear km treated and targets and traps used detailed in S1.Annex 2.

These financial costs represent the monetary expenditures by PNLTHA directly associated with the implementation of vector control. They encompass the tangible, measurable, and explicit financial outlays required for the implementation of the activity. In contrast, economic costs go beyond the explicit financial expenses and consider opportunity costs— the value of resources that could have been used elsewhere but were allocated to vector control and the value of unpaid inputs such as donated drugs or targets or unpaid local community labour [33]. In this provider’s perspective study, as PNLTHA paid for all staff, vehicles and other inputs into the control activity, there were not too many adjustments to be made to convert from the financial to the economic viewpoint. For example, in this study both staff and vehicles were employed full-time on the VC work, and none were shared with other activities or were borrowed from other organisations as was the case in some of the recent HAT vector control activities in other countries [31, 32]. However, some of the management costs benefitted other activities.

The study considered resources with a useful life of less than 12 months as recurrent costs and resources with a useful life of over a year as capital costs [34]. For the economic analysis, the capital costs were annualized using straight line depreciation and assuming they had no residual value at the end of this period. Thus their purchase value was divided by their lifespan. Useful life estimates were based on discussions with experts and on WHO-Choice guidelines [35].

A mixed-methods approach was used to estimate the annual costs to treat 210 linear km and to protect 1,925 km², this being the ultimate extent of the whole operation after 5 deployments (Figure 1 and S1.Annexe II). Although PNLTHA was the sole implementer, costs were recorded in different locations and at different levels of the organization. Thus, bottom-up micro-costing was used to estimate costs that were directly allocated to target deployment in the field. We collected detailed data on the quantities of inputs and prices to value the resources used. Additionally, we used a step-down or gross costing approach to estimate costs that could not directly be attributed to specific field activities, such as management and transport costs [36]. The current awareness-building and management strategy were costed using information from current activities (2018-2019), as a standardized strategy for these activities was only implemented after the study period. The management support unit at the provincial level accompanied the vector control activities in four health districts, including Yasa Bonga. Therefore, in the economic analysis, the annual cost of management support was divided by four to calculate the management costs attributable to the health district of Yasa Bonga.

A 5-year period was selected for estimating the financial costs for treating the whole area, with the aim of showing what the provider’s costs over time would be for funding such an operation. The time period chosen reflected that capital equipment with the highest cost, namely the vehicles, was determined to have the longest estimated lifespan of 5 years according to discussions with field experts. Thus, we took the costs collected for the actual operation and put them together to show how a 5-year project would look. Then, we made the appropriate adjustments for converting them to economic costs as explained above, replacing capital costs with annual depreciation and adjusting the share of management costs. These costs were not discounted as we only looked at the cost of one year of vector control deployment.

Afterwards, the results were combined to calculate the total annual cost to implement tsetse control covering the entire health district and the cost per activity, namely entomological monitoring and surveillance, sensitization, biannual target deployment, and management support costs. The costs were presented in four main categories: human resources (HR), transport-related costs (fuel, vehicle use and maintenance, and rent of pirogue), specialized equipment, and other (stationary, small camping equipment, etc.) [17, 31, 32]. Then we looked at the main cost drivers, namely specific expenses that significantly influenced or contributed to the overall costs.

In order to be able to compare the results from Yasa Bonga with gHAT vector control using Tiny Targets in other settings, the cost was also expressed as a cost per (i) area protected (USD/km²), (ii) cost per target used (USD/target - accounting for the number of targets used annually to protect the area), (iii) target deployed (USD/target - factoring in the number of targets deployed in the protected area), (iv) length of river controlled (USD/km), and person protected (USD/person). We performed a univariate sensitivity analysis for the economic cost of the main cost drivers to evaluate the impact of these drivers on the overall costs. This analysis included considering annual and triannual sensitization, varying the transport costs (+-10%), including the cost of the vector control unit at central level and varying the number of health districts management by a vector control unit at provincial level. Lastly, the results were compared with cost estimates of vector control with Tiny Targets in other settings.

All costs were recorded in the currency in which they were incurred and converted to USD using the average exchange rate between January 2015 and September 2017, which were Euro to dollar: 1.13 and Congolese franc (CDF) to dollar: 0.00084.

## Results

Over this 5-year period, the projected annual financial costs for covering the whole area protected by the end of 2017 would thus range from 104,630 USD to 168,478 USD, with the highest cost in the first year reflecting the initial capital investments required, as illustrated in Table 1. Sensitization and management support costs would be highest in the first year, while costs associated with monitoring, surveillance, and target deployment remained consistent across the years. The number of targets deployed increased between 2015 and 2017, correlating with the expanded linear coverage, as illustrated in Figure 1. For a detailed breakdown of yearly costs during the scale-up, refer to S1.Annex III. The overall financial cost over the 5-year period is projected to decrease by approximately 113,000 USD or almost 20% if the targets are donated by the manufacturer.

As illustrated in Table 2, the total average annual economic cost for vector control in the health district of Yasa Bonga to treat 210 linear km is 120,127 USD. Almost 50% of the cost is linked to the biannual target deployment, and the management and sensitization each represent around 20% of the costs (see Figure 2). The cost per target used was estimated at 5.31 USD, 573 USD per linear km of river treated, or 62.4 USD per km^2^ protected. Costs that could not directly be attributed to a specific category were reported under “Other,” such as glue to assemble the targets, camping equipment, phone, and internet credit. How financial costs were converted into economic costs is shown in S2.Supplementary Spreadsheet and S1.Annex III also details the cost per deployment and monitoring and surveillance round.

**Figure 2.**
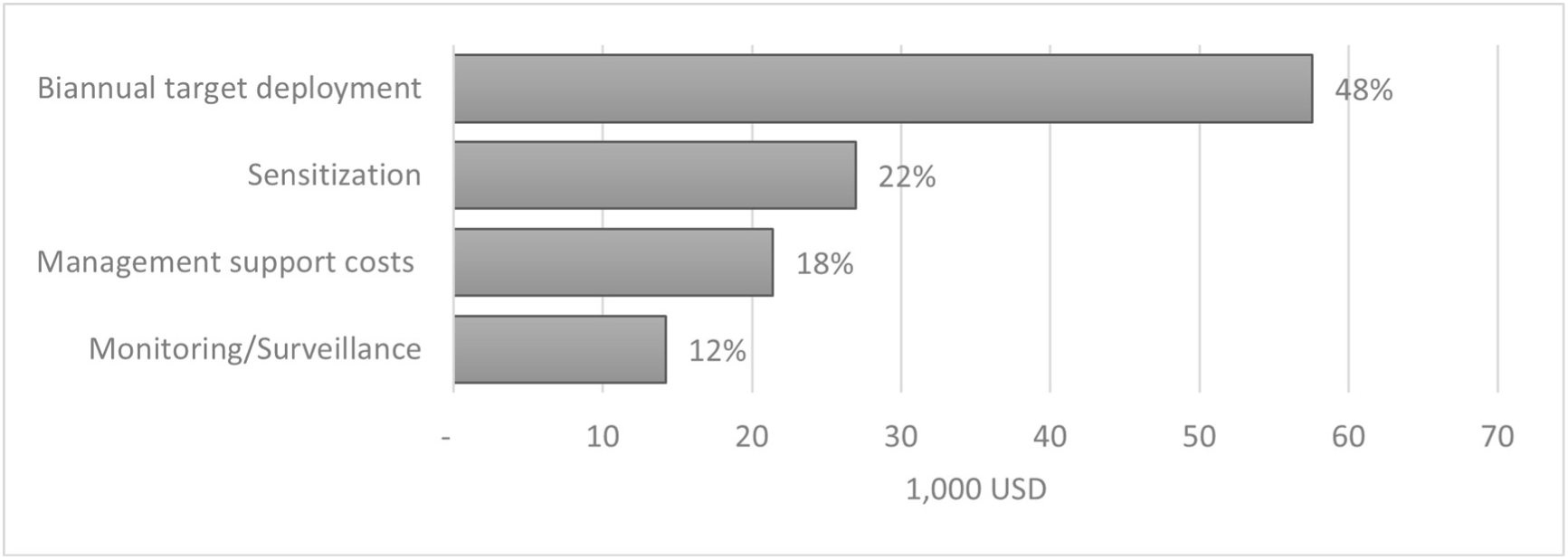
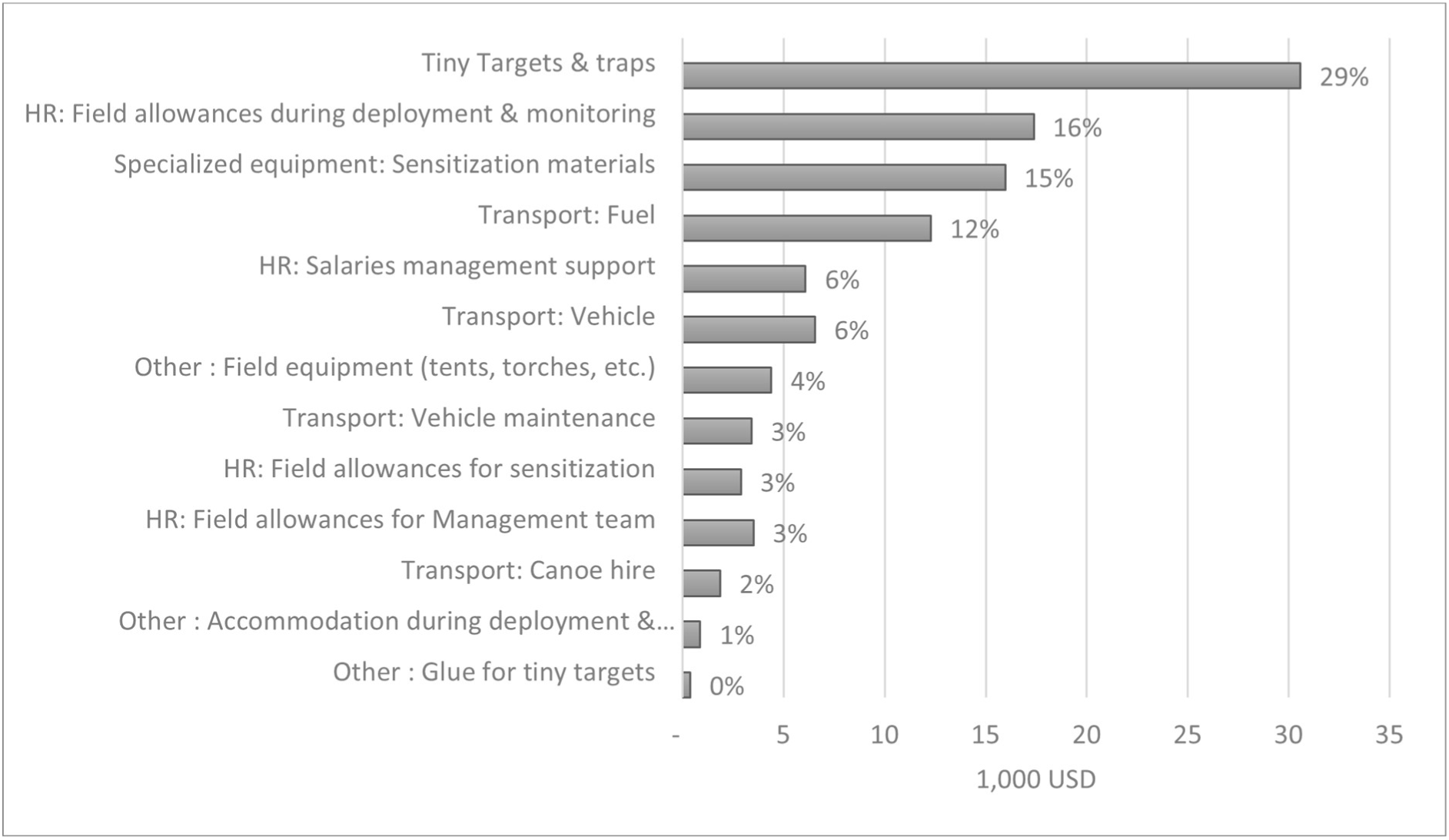
Contribution of the different activities to the overall annual economic cost (in 1,000 USD)

**Table 2.** Estimated average annual economic cost to cover 210 linear km based on the costs reported between 2015 and 2017 (USD). Calculations are shown in the S2.Supplementary Spreadsheet.

| **Activity/**<br>**Cost category** | **Total**<br>**USD** | **%** |
| --- | --- | --- |
| **Monitoring/Surveillance** | **14,214** | **12** |
| HR | 2,610 | 2 |
| Transport | 5,443 | 5 |
| Traps | 4,189 | 3 |
| Other | 1,972 | 2 |
| **Sensitization** | **26,929** | **22** |
| HR | 2,888 | 2 |
| Transport | - | 0 |
| Specialized equipment | 15,964 | 13 |
| Other | 8,077 | 7 |
| **Biannual target deployment** | **57,571** | **48** |
| Tiny Targets | 26,399 | 22 |
| HR | 14,016 | 12 |
| Transport | 10,198 | 8 |
| Other | 6,959 | 6 |
| **Management support costs** | **21,413** | **18** |
| HR | 10,345 | 9 |
| Transport | 9,035 | 8 |
| Specialized equipment | - | 0 |
| Other | 657 | 1 |
| Trainings/meetings | 1,317 | 1 |
| **Total cost** | **120,127** | **100** |
| **Cost per target used (22,622)** | **5.31** |  |
| **Cost per target deployed (11,311)** | **10.62** |  |
| **Cost per linear km (210)** | **573** |  |
| **Cost per km<sup>2</sup> protected (1,925)** | **62.4** |  |

The main cost drivers are the purchase and import of Tiny Targets and traps (29%), human resources during the deployment and monitoring (16%), sensitization equipment (15%) and fuel (12%) (Figure 3). This is also reflected in the breakdown of the costs per activity as shown in Figure 2.

**Figure 3.**
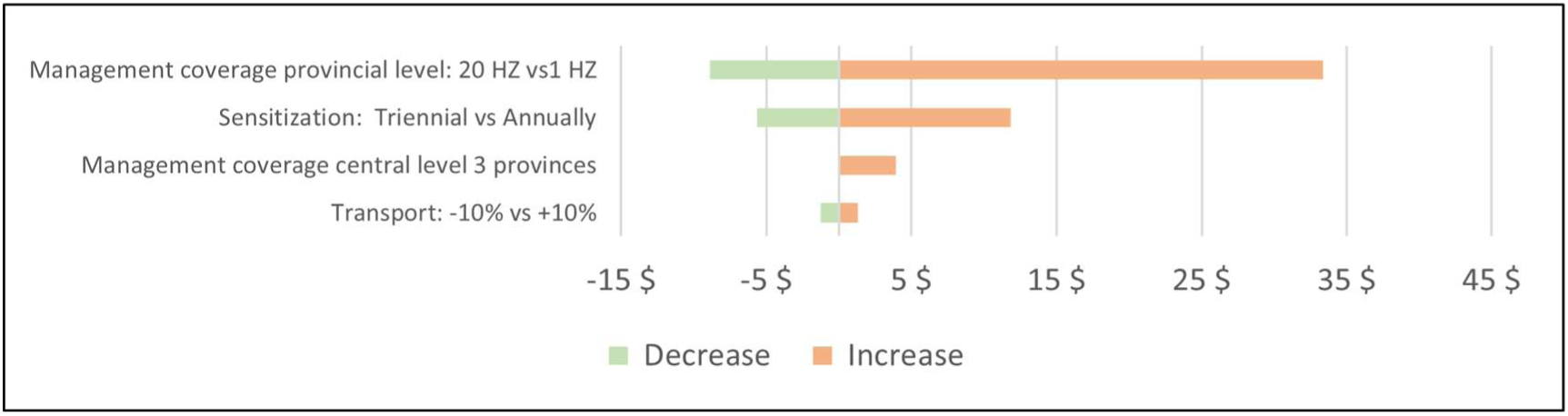
Sensitivity analysis – Economic cost per km² protected

### Sensitivity analysis

The sensitivity analysis looked at changes in the main cost drivers on the economic cost per km² and the economic cost per linear km of vector control deployed (Figures 4-5). Changes in the transport cost had a minor impact on the overall cost. The cost would decrease by 10% if the sensitisation would take place once every three years instead of biannually. The most significant impact on the cost can be seen in the coverage of the management unit, namely the number of health districts the current set-up of the management unit could accompany and supervise. Setting up such a unit for a limited number of health districts would drastically increase the overall cost of vector control. On the other hand, the cost would decrease if health districts could independently deploy vector control with limited strategic support from the provincial level so these resources could be used to cover a larger area but this could also negatively impact the quality and traceability of the intervention. If a provincial unit were able to cover 20 health districts the overall cost would decrease with 55%.

**Figure 4.**
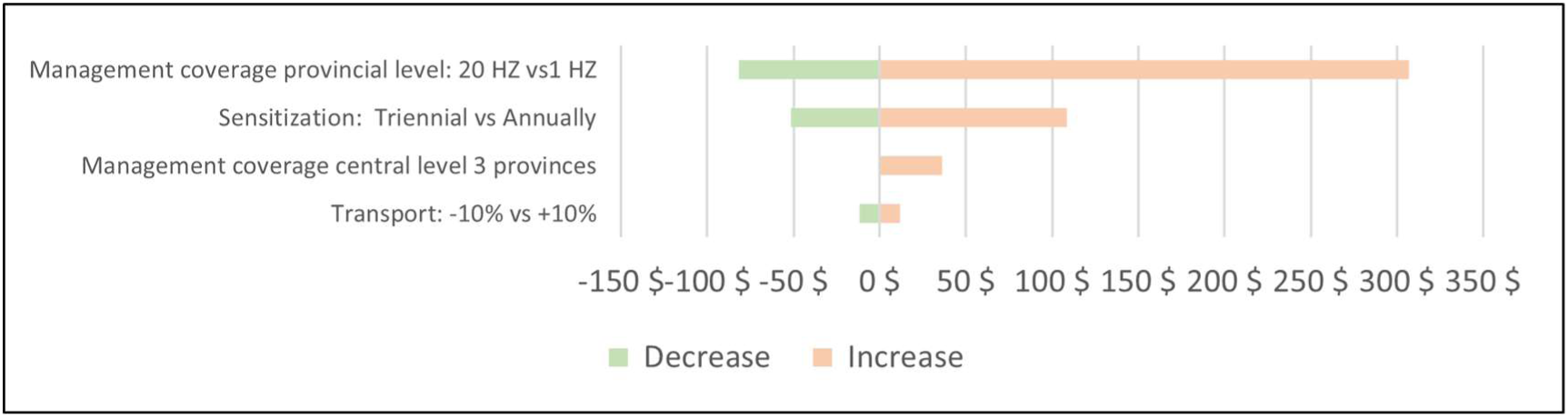
Sensitivity analysis – Economic cost per linear km

Furthermore, the estimated cost did not take into account the cost of the vector control management unit at central level that supports 11 health districts in 2 provinces. Including this cost would increase the overall cost by approximately 6%.

### Comparison of cost of HAT vector control using Tiny Targets in other settings

Lastly, in Table 3, we look at the cost of the work in Yasa Bonga, compared to the costs calculated for the three other Tiny Target costed [17, 31, 32]. The cost comparisons vary greatly depending on which criterion used. The annual cost per target deployed is much lower than in other countries, while the cost per area controlled (USD/km²) is similar to Arua, Uganda, and Mandoul, Chad. A comparable cost to Chad was observed for the cost per km² protected. Still, since Yasa Bonga is more densely populated than Mandoul, the intervention was cheaper regarding cost per person protected, yielding a value similar to that in Côte d’Ivoire. One of the reasons for the lower annual cost per target deployed in Yasa Bonga and Uganda, compared to Chad and Côte d’Ivoire is due to the organizational context. Activities in the DRC are done by locally recruited staff supervised by provincial management teams resulting in lower salary costs and lower costs related to supervision visits (per diem, transport, etc.)

**Table 3.** Comparison of annual vector control activities and costs in different settings.

| Description<br>(costs in USD) | Chad | Côte<br>D'Ivoire | Uganda | DRC |
| --- | --- | --- | --- | --- |
|  | 2015-2016 | 2016-2017<sup>1</sup> | 2012-2013<sup>2</sup> | 2015-2017<sup>3</sup> |
|  | [21, 31, 32] | [19, 31] | [17, 18, 31] | [13] |
| **Annual economic cost** | **56,133** | **61,253** | **21,982** | **120,127** |
| Number of targets deployed<br>in the area covered | 2,708 | 1,939 | 1,551 | 11,311 |
| Target maintenance | Annual<br>redeployment | Annual<br>redeployment | 60%<br>redeployed in<br>2012/13 | Biannual<br>redeployment |
| Annual number of targets<br>used in the covered area | 2,708 | 1,939 | 2,501 | 22,622 |
| **Annual cost per target**<br>**deployed in the area**<br>**covered** | **21** | **32** | **14** | **11** |
| Number of linear km treated | 45 | NA | 78 | 210 |
| Targets per linear km | 60 | NA | 20 | 54 |
| **Annual cost per linear km**<br>**treated** | **1,244** | **NA** | **283** | **573** |
| Number of km<sup>2</sup> treated | 45 | 130 | 16 | NA |
| Targets per km<sup>2</sup> treated | 60 | 15 | 97 | NA |
| **Annual cost per km<sup>2</sup>**<br>**treated** | **1,247** | **471** | **1,374** | **NA** |
| Number of km<sup>2</sup> protected | 840 | 130 | 250 | 1,925 |
| Targets per km<sup>2</sup> protected | 3 | 15 | 6 | 6 |

| **Annual cost per km<sup>2</sup>**<br>**protected** | **67** | **471** | **88** | **62** |
| --- | --- | --- | --- | --- |
| Number of people protected | 39,000 | 120,000 | 112,500 | 165,062 |
| Population density<br>(person/km<sup>2</sup> protected) | 46 | 923 | 400-500 | 86 |
| **Annual cost per person**<br>**protected** | **1.44** | **0.51** | **0.20** | **0.73** |

<sup>1</sup> NA – Not applicable as Tiny Targets were deployed throughout the intervention area.
<sup>2</sup> Costs were converted to 2016 prices [31]. Half way through year 2012-13 for which the costs were analysed, about 60% of targets were replaced. In subsequent years, Uganda moved to biannual redeployment as in DRC. The number of people protected in Uganda was estimated at between 100,000 and 125,000.
<sup>3</sup> NA – Not applicable as Tiny Targets were deployed on the narrow fringing vegetation of the rivers.

## Discussion

This study estimates the costs of vector control for HAT using Tiny Targets in the DRC. At a total economic cost of 120,127 USD or a financial cost ranging between 104,630 USD and 168,478 USD per annum for protecting an area of 1,925 km, depending on the assumptions made, the cost per person protected comes to 0.73 USD or 11 USD per target deployed. At the beginning of the Tiny Target operation, financial costs are driven by sensitization and management, after which target deployment dominates the costs. In terms of the average annual economic costs, almost 50% are attributed to deployment, and 20% each to management and sensitization. Monitoring and surveillance account for the lowest proportion of costs, both in economic and financial terms. Currently the Tiny Targets are donated by the manufacturer which is projected to reduce the financial cost by almost 20%.

In this paper we have emphasized both the costing methodology and the nature and detail of the information required. Our objective was to help enable such cost analyses of vector control work to be conducted in other settings, not just retrospectively, at the evaluation stage but also before work is undertaken, when first planning an intervention. Thus, S1.Annexes contains not just details supporting the calculations presented above, but also of the individual components of the deployment, trap monitoring and sensitisation costs. More information is available in the S2.Supplementary spreadsheet. The basic methodology of collecting financial costs and adjusting these to better reflect the total economic or societal cost has been explored to some extent in the previous papers on the cost of Tiny Target operations to control HAT transmission but here we emphasize and contrast the two approaches [17, 31, 32].

Assessing the cost-effectiveness of tsetse control is challenging due to the complexity of factors influencing vector control costs and effects. A comparison of results for the DRC with those from other countries suggests significant variations in vector control costs. In HAT endemic foci, adapting vector control strategies to the local context is essential, introducing variability in the overall costs. Numerous factors contribute to this variability, including diverse tsetse habitats such as expansive forests, mangroves, swamps, and narrow linear riverbanks. Further the choice of vector control strategy hinges on considerations such as target coverage, deployment methods (ranging from canoes and on-foot approaches to the use of cars and motorbikes), the availability of local manpower and the frequency of target deployment and monitoring. All these factors contribute to the intricacies of the overall cost.

Defining the effectiveness of vector control for sleeping sickness involves evaluating its impact on reducing disease transmission by tsetse flies, primarily with the preventive objective of mitigating and ideally halting HAT transmission. In contrast, other HAT control measures, namely case detection and subsequent treatment, operate reactively by addressing cases post-infection. Vector control efficacy could be measured by the reduction in cases or the number of cases averted. The prevalence of the disease within a protected area will significantly influence the effectiveness, measured in terms of cases averted or Disability-Adjusted Life Years (DALYs) avoided.

Directly comparing the cost-effectiveness of a vector control intervention is challenging, as it is seldom the only control measure implemented and it’s difficult to measure it’s impact on transmission. Therefore, the cost-effectiveness of vector control is typically expressed in metrics like cost per km treated, cost per km² protected, or cost per population protected (or cost per case averted when feasible). However, these indicators are influenced by factors external to the vector control intervention, such as baseline prevalence, population density in the protected area, or the size of the protected area compared to the treated area. Therefore, it is crucial to consider the denominator when interpreting the cost-effectiveness indicators presented in this paper. Vector control with Tiny Targets appears to be more cost-effective in terms of cost per person protected in Uganda, Côte d’Ivoire, and the DRC as the tool was deployed in an area with a higher population density, resulting in a relatively lower cost per person protected than in Chad [17, 31, 32].

For sleeping sickness, an optimal strategy would likely integrate case detection initiatives aimed at promptly identifying and treating infected individuals with vector control methods targeting the reduction of tsetse fly transmission. The collaborative impact of these approaches synergizes to most likely yield a substantial reduction in sleeping sickness cases. The evaluation of their individual and combined effectiveness and costs is the focus of the Human African Trypanosomiasis Modeling and Economic Predictions for Policy (HAT MEPP) project [37].

While several studies demonstrated that vector control of tsetse flies can play an essential role in HAT elimination, using Tiny Targets presents several limitations and challenges. Significantly reducing the HAT disease burden through tsetse control relies on its ability to reduce transmission effectively. HAT tends to persist in remote and rural areas with dense vegetation near water sources such as rivers, lakes, and ponds. The lack of comprehensive, accurate geospatial data on tsetse fly habitats, along with limited information about the actual “transmission zones,” the sites where people get bitten by infected tsetse, makes it challenging to identify locations where vector control could impact transmission. This requires geospatial modelling followed by on-site entomological surveys, but limited information is available on the complete actual cost of these preliminary evaluations [17, 30–32]. These knowledge gaps also make it challenging to develop a uniform vector control strategy or to determine the necessary “quantity” of vector control per square kilometer needed to halt transmission throughout the countries affected by the disease [19, 38].

Additionally, an exploratory entomological survey in Yasa Bonga conducted in the same period as the study on community-based control of tsetse (’17-’18) revealed that fishponds provided suitable habitats for tsetse. Nevertheless, covering these areas with large-scale vector control interventions led by individuals not familiar with the local environment is challenging due to the difficulty in locating and navigating such areas, which would significantly increase the workload of the vector control teams. A study in the southwest of the health district (see Figure 1) showed that these areas could be covered with a community-based approach to Tiny Targets deployment, organized and managed by local community members. However, data available on the costs for this type of intervention is currently unavailable [39].

Currently, the DRC vector control for sleeping sickness is successfully being implemented by local teams managed by the PNLTHA. This is taking place in a select number of health districts located in provinces adjacent to the Kinshasa province and accessible by car from the capital. The activities were successfully continued during the COVID-19 pandemic, which shows that a sustainable system was developed that transitioned responsibilities to provincial and health district levels [40]. Introducing vector control activities in remote and resource-constrained areas in other provinces of the DRC will require additional investment for the preliminary geospatial and entomological studies needed, the creation of vector control capacity (resource, equipment, and trained personnel, etc.) and higher transport costs, compared to Bandundu, due to the distances and a higher fuel cost. Initial costs related to training, infrastructure development, and equipment acquisition would be spread over a shorter duration if the intervention would be implement for a limited period due to the context of disease elimination resulting in a higher average cost. HAT vector control might leverage an existing supply chain and management system by integrating this activity into the broader health system while reinforcing the entire health system beyond this specific disease focus.

Vector control using Tiny Targets has proven to be a feasible tool at a lower cost than former methods [41]. Therefore, Tiny Targets can play an important role in the HAT elimination strategy as it could help stop transmission in foci where the disease persists. The successful scale-up of a Tiny Targets will require a good local understanding of the terrestrial and aquatic ecosystem of tsetse fly habitats and the development of tsetse control measures where provincial and health district levels play an important role. The implementation cost of this approach can be drastically reduced when implemented on a large scale, with a local vector control management unit covering a larger geographical area for minimum duration of 5 years, allowing full use of the investments needed to build local capacity and awareness. While any measure aimed at elimination will present some challenges, this study, like other costing studies on Tiny Targets, shows that the cost can be quite accessible.

## Data Availability

Data available in supporting files

## Acknowledgments

The authors acknowledge the staff of the PNLTHA and the fieldworkers in Yasa Bonga for their contributions under arduous field conditions. The authors also would like to thank Professor Stephen Torr and Dr. Andrew Hope for their valuable contributions to the final draft of the article.

